# Supporting Families of Children with Cancer: Development and Evaluation of a Virtual Assistant for Social Care

**DOI:** 10.64898/2025.12.03.25341557

**Authors:** Emre Sezgin, Adelaide M. Booze, Lydia M. Wisne, Daniel I. Jackson, Micah A. Skeens

**Affiliations:** The Abigail Wexner Research Institute at Nationwide Children’s Hospital, Columbus OH; The Ohio State University College of Medicine, Columbus OH

## Abstract

**Purpose:** Families of children with cancer face substantial financial, social, and emotional stressors, including unmet health-related social needs (HRSN) and toxic stress, which contribute to disparities. Virtual assistants (VAs) offer a scalable option to screen for HRSN and connect families to resources, but their design and acceptability in pediatric oncology are not well characterized.

**Methods:** A two-phase mixed-methods study was conducted to co-design and evaluate a VA for caregivers of children with cancer. In Phase 1, a community advisory board including healthcare providers and a parent advocate (CAB; *N*=8) iteratively refined VA content, tone, and interface through three virtual co-design sessions. In Phase 2, caregivers (*N*=40) completed in-person VA prototype usability sessions that included structured observation, a semi-structured interview, the System Usability Scale (SUS), caregiver satisfaction survey, and a reaction card task. Social drivers of health, rurality, and neighborhood disadvantage were characterized using survey data and geocoded indices.

**Results:** CAB feedback refined prototype components including community resources, stress management, conversation history, and resource feedback. In Phase 2, usability was high (SUS *M*=82.6, *SD*=14.0); 95% of caregivers rated the VA easy to use, good to use, and most reported the VA motivated them to stay engaged with screening and resources. Reaction card responses were predominantly positive (e.g., “accessible,” “easy,” “useful”). Higher social vulnerability was modestly associated with higher usability scores, while rural and medically underserved caregivers reported lower confidence and more negative desirability attributes.

**Conclusion:** The stakeholder-informed VA to address HRSN and toxic stress in pediatric oncology was feasible, highly usable, and acceptable. Future work should test impact on toxic stress, service uptake, and equity in larger, more diverse and longitudinal studies.

## INTRODUCTION

Pediatric cancer is a leading cause of disease-related death among children, and while survival rates have improved, the incidence of pediatric cancer continues to rise with persistent disparities in outcomes.^1,2^ There is an emerging need to provide social care support for those with high needs. Within social care, specifically Health related Social needs (HRSN), including financial, nutritional, and environmental support, can impact the ability of children and their families to cope with disease and access healthcare services.^33^ Furthermore, living in impoverished and unhealthy conditions puts children and families under chronic stress, which can overpower their stress response systems. This condition, known as “toxic stress”, has been associated with unmet HRSN and negatively impacts a child’s brain structure and raises the likelihood of experiencing various health issues, including physical, behavioral, socio-emotional, and cognitive problems.^4,5^ Therefore, providing social care is essential, including screening for unmet HRSN, social drivers of health (SDOH) and related mental health needs for identifying and addressing these needs, reducing health disparities and improving quality of life.^6^

Traditional screening methods for social needs are often burdensome, resource-heavy, and may not provide access to accurate timely information or connect with professional care providers, especially in low resource clinics.^7,8^ In such cases, digital health technologies have shown promise in supplementing healthcare related services.^9–11^ At clinics, electronic health records (EHR)based and tablet PC facilitated SDOH screenings have been employed to support routine screening, and offload social work efforts.^9,12^ In addition, virtual assistants (also known as conversational agents and chatbots) have been emerging for healthcare communications including mental health screening and social support^13,14^ and community resource access.^15,16^ Chatbots provide scalable, personalized, and user-friendly platforms for patient interaction and data collection.^17–20^

This study builds upon the foundational work of a virtual assistant (VA) designed to provide social care support, specifically to screen for unmet HRSN and guide users to community-based resources.^16^ The current work expands this effort through stakeholder-informed co-design and evaluation in a pediatric oncology context. The primary goal was to adapt and refine the VA and evaluate its usability and acceptability among caregivers of children with cancer. Our central research questions were: “How can a user-centered VA improve HRSN and stress outcomes for families of children with cancer?” and “What are key factors influencing its feasibility and acceptability?”

### Objectives

The overarching goal of this study was to adapt and evaluate a VA to screen for SDOH and reduce toxic stress among families of children with cancer. Specifically, the study aims to (1) co-design (refine) the VA with input from key stakeholders and (2) assess its usability and acceptability among caregivers.

## METHODS

This study consisted of two phases: (1) co-designing VA with caregivers of children with cancer through a community advisory board (CAB) and employing a human-centered iterative design approach, and (2) investigating VA feasibility and acceptability through mixed methods user testing including system usability scale and qualitative interviews.

### Phase 1 Study Design

Phase 1 consisted of three co-design and development sessions with a CAB, which included participants from a large free-standing children’s hospital and a patient advocate. All CAB members were 18 years old or older and provided informed consent prior to participation. Each session was held virtually lasting an average of 46 minutes. During the virtual session, CAB members reviewed the evolving prototype with prior feedback incorporated. This process continued until consensus was reached that it was ready for phase 2 testing.

**Figure 1.**
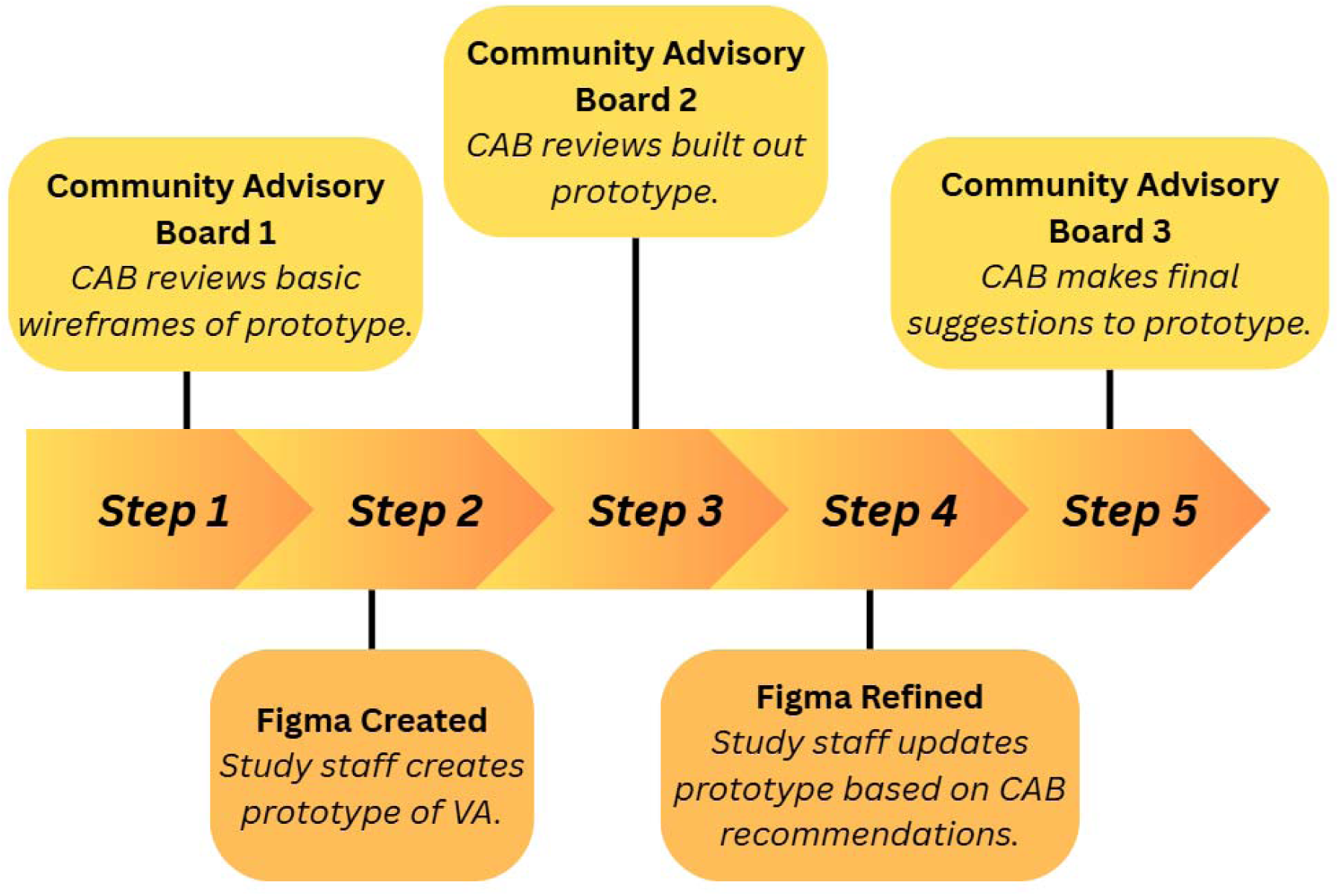
Phase 1 community advisory board session progression.

Participants in the initial CAB were presented with wireframes of the VA in an app format via Miro, a collaborative design platform. The study team adopted an iterative, co-design approach, incorporating changes based on CAB member suggestions. Following the first CAB, Figma, a digital interactive prototyping tool, was used to implement feedback. By the third CAB session, members largely approved the VA and its features, deeming the prototype suitable for presentation to families in Phase 2.

### Phase 2 Study Design

Phase 2 involved a cross-sectional mixed-methods evaluation of the VA. Caregivers were recruited in person from a large Midwestern children’s hospital between October 2024 and February 2025. Eligible caregivers were identified from the Hematology and Oncology clinic schedule based on the following criteria: (1) age 18 years or older, (2) English-speaking, (3) having a child aged 0-17 years with a cancer diagnosis, and (4) whose child was actively receiving cancer therapy at recruitment and during the study period. Of the 57 eligible caregivers approached, 41 consented to participate, while 16 declined due to reasons such as lack of time, lack of interest, discomfort with research, or a preference for virtual participation. One caregiver subsequently withdrew, resulting in a final cohort of 40 caregivers. Quantitative and qualitative data were collected in person to obtain feedback on the VA’s interface, usability, satisfaction, reactions, and content. Caregivers were evaluated for VA engagement and understanding through the following steps:

Step 1 – Unobtrusive observation to measure the intuitiveness of the interface (10 - 15 minutes): Caregivers were briefly introduced to the concept of the VA and asked to start interacting with it. The study team did not intervene or provide instruction but observed and recorded participants’ progress.

Step 2 – Interactive Observation (10-15 minutes): The study team began to interrupt participants during their interaction with the VA by posing questions regarding observed cues that implied success or delay in program use. Caregivers were encouraged to verbalize their criticisms of the VA. Since the discussion was based on evolving patterns of interaction, pre-prescribed questions were not possible.

Step 3 – Debriefing (15-20 minutes): Caregivers were asked to reflect on their experience with the VA, its interface and contents, and share their opinions on the strengths and weaknesses. At Step 3 conclusion, caregivers completed a 15-minute semi-structured interview regarding the VA. They were asked about their initial impressions, perceptions of similar tools, feature preferences, perceived helpfulness, missing elements, suggestions, and potential barriers or facilitators of use for their family. Following the interview, caregivers completed the System Usability Scale (SUS, assessing functionality and acceptability), Caregiver Satisfaction survey (assessing satisfaction), and Reaction Card (assessing desirability).

### Measures

During Phase 1, we collected limited personal data. Other than qualitative input, CAB members reported on their sex, race, ethnicity, age, relationship status, education, employment status, occupation and work setting.

The following measures were collected only from phase 2 participants.

#### Demographic Characteristics

Caregivers provided comparable sociodemographic information (sex, race, ethnicity, relationship status, education, employment status, occupation and work setting, household composition, family income, and spouse’s educational attainment.), as well as additional information regarding their relationship to the child and the child’s age, sex, race, ethnicity, educational status, and grade level. Information about child diagnosis and treatment were obtained from EHRs.

#### Functional Impact of Toxic Stress for Parents (FITS-P)

Caregivers completed the four-item Functioning in Treatment Scale-Parent version (FITS-P),^21^ reporting dichotomously (yes/no) on each item. Items assessed caregiver difficulty in the past month with (1) managing thinking (e.g., intrusive thoughts, poor concentration), (2) regulating behavior (e.g., irritability, difficulty with tasks), (3) maintaining schedules (e.g., attending work or appointments, managing child care), and (4) managing the caregiver-child relationship (e.g., reduced enjoyment, frustration in caregiving).

#### PhenX Toolbox

Caregivers completed the PhenX Measures for SDOH. This instrument provides standardized measurement protocols intended to facilitate the development of effective interventions for mitigating health disparities.^22^ The specific measures completed by caregivers include current address, health insurance coverage, food insecurity, health literacy, access to health services, internet access, access to health technology, and perceptions of housing insecurity.^23^

#### System Usability Scale

Caregivers completed the System Usability Scale, a 10-item questionnaire routinely employed to assess the functionality and acceptability of digital health applications.^24^Items are rated on a 5-point scale, with scores ranging from 0 to 100. The scale’s reliability (Cronbach’s alpha= 0.91) and validity (Pearson correlation r= 0.81 with a 7-point scale of “user-friendliness”) are well-established.^25^ A score exceeding 68 is considered above average.^26^

#### Caregiver Satisfaction

Caregiver Satisfaction was evaluated utilizing a 9-item survey, developed by the investigators, to ascertain caregiver perspectives on the VA. This included perceived benefits, burdens, barriers, suggested modifications, and overall satisfaction. Items were scored on a 4-point scale, with higher total scores indicative of greater satisfaction.^27^

#### Reaction Card

A reaction card methodology was employed to assess the desirability of the Virtual Assistant (VA) based on user experience. Developed by Microsoft as part of a “desirability toolkit,” reaction cards are designed to elicit user reactions, thoughts, or opinions regarding a specific tool or technology.^28^ Caregivers provided feedback on VA desirability by utilizing a reaction card featuring 55 pre-listed words. The frequency with which each word was selected was subsequently summarized to indicate the overall attitude toward the VA. A higher incidence of positive words suggests greater usability, whereas a higher incidence of negative words indicates lower usability.^29^

#### Rural Residency

Caregivers’ addresses were categorized as rural or non-rural based on Rural-Urban Commuting Area (RUCA) codes. These codes, derived from census tract and ZIP-code level data, quantify population density, urbanization, and commuting patterns on a scale of 1 to 10. In alignment with established research, rural residency was defined by scores of 4-10 (micropolitan to rural), while non-rural residency encompassed scores of 1-3 (metropolitan).^30,31^

#### Social Drivers of Health

Participant addresses were coded for SDoH to inform if non-medical factors associated with caregiver residency influence VA usability. The CDC Agency for Toxic Substances and Disease Registry developed the Social Vulnerability Index (SVI), which uses 16 variables from the U.S. Census American Community Survey to identify communities who may need support before, during, or after disasters.^32^ These variables are grouped into four themes: socioeconomic status, household characteristics, racial/ethnic minority status, and housing type and transportation, and can be grouped into a single overall social vulnerability measure. SVI scores range continuously from 0 and 1, with higher scores indicating greater vulnerability. Medically Underserved Area (MUA) designations are based on geographic location and used to identify access to healthcare services, where a 0 represents medically served, and 1 represents medically underserved.^33^ Finally, Appalachian residency is determined by county, referencing lists established by the Appalachian Regional Commission (ARC), an organization who identifies and helps support the economic development in Appalachian areas.^34^ Addresses were coded as 0 for a non-Appalachian county and 1 for an Appalachian county.

### Virtual Assistant Prototype

The conceptual framework for our VA was derived from our previous work on an HRSN chatbot designed for pediatric primary care, serving as an exemplar for the VA’s conversational architecture, organization, and subject matter. Subsequent feedback obtained from a CAB guided the development of the VA prototype. Following this, CAB members evaluated the prototype and granted approval for its assessment by caregivers. Figure 2 outlines the wireframes on how we started with CAB 1 and iteratively improved and refined after CAB 2 and 3, finally used for phase 2. **(See supplementary materials 1 for details on iterative design updates).**The VA comprised four main functionalities:(1) a Community Resource module (2) a Stress Management module (3) a History tab (4) a Feedback loop.

The community resource module began with an introduction from the VA and asked if caregivers were looking for food, housing, transportation, or financial resources. After selecting a resource category, caregivers complete a screening section with pre-populated answers to simulate the VA’s functionality. Screening questions included location (Zipcode), dependents’ ages, and child health conditions. Based on responses, the VA provided a mock-up resource that the caregiver could explore in detail. Resource features included the option to “save” the resource, and review eligibility criteria, required documents, benefit program options, application guidance, and other additional information (i.e., address, business hours, cost, phone number, documents, eligibility, and which benefits can they use). More resource details such as contact information, a “meet the team” feature and map links were also available.

The Stress Management module asked how caregivers were feeling that day, then asked if they felt stressed that day. If they said yes, the FITS-P informed questions were asked for evaluating difficulties with managing thoughts, behaviors, schedules, and relationships with children. At the end of the screener, resources such as crisis text-line information and optional mindfulness exercises were offered. Throughout, Caregivers could access explanations of terms and questions via “learn more” prompts (e.g., “learn more” in regards to has it been hard/difficult to manage your behavior would supply a bubble with “such as: losing patience easily, saying things you quickly regret, can’t bring yourself to do your daily tasks). The mindfulness exercise guided caregivers through progressive muscle tightening and a breathing circle, supported by visual instructions.

**Figure 2.**
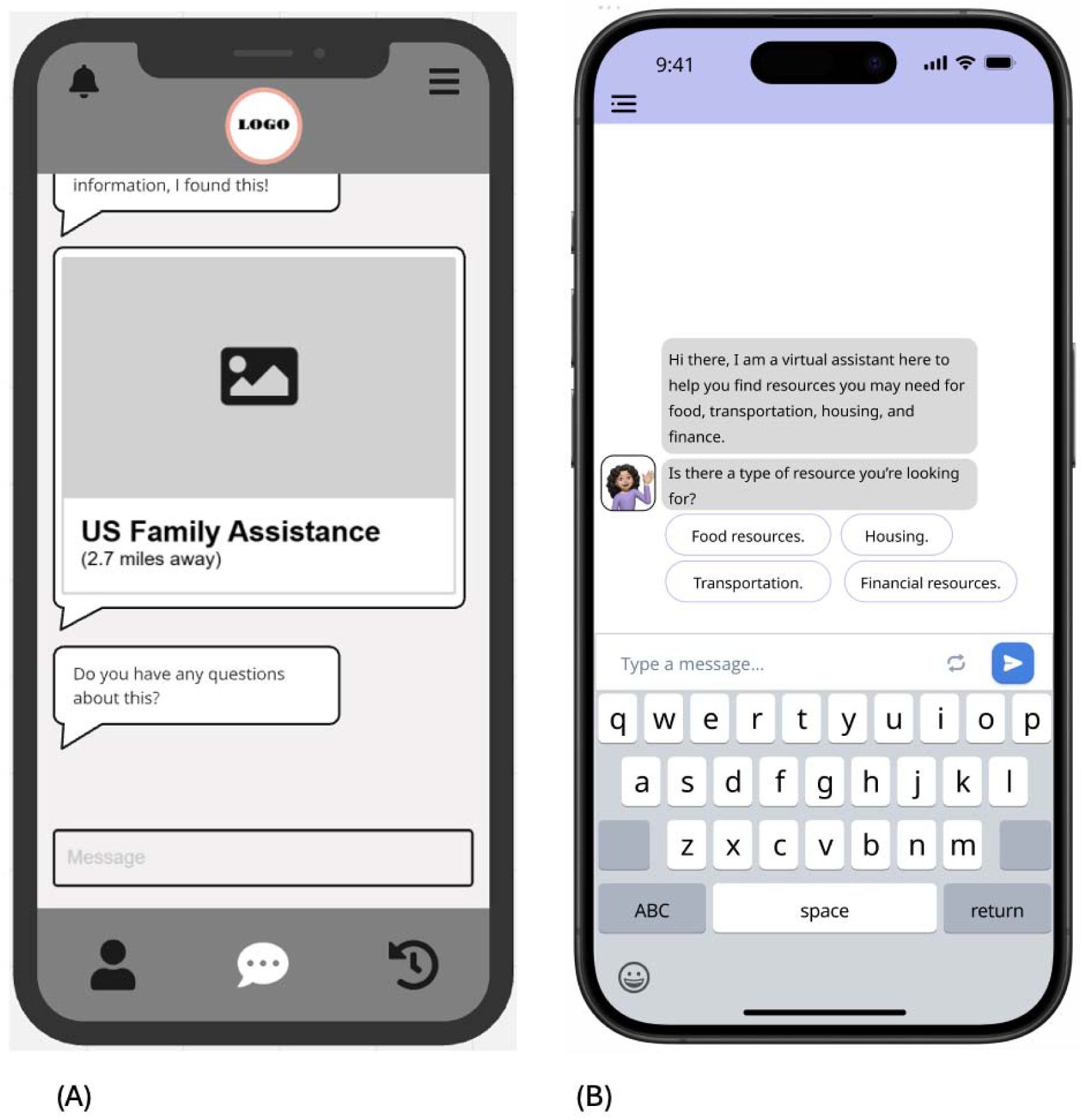
*Example wireframe (A) and prototype (B) visuals*

### Statistical analysis

Data were cleaned and analyzed using IBM SPSS, version 28 for Windows. For the CAB, descriptive statistics (frequencies, means, percentages, *SD*) summarized demographic data. Semi-structured interviews were transcribed and then used rapid analyses to extract major themes.^35^ For Phase 2, descriptive statistics, bivariate correlations, and *t*-tests summarized quantitative data from surveys. Semi-structured interviews were transcribed verbatim for content analysis and coded using NVivo software. Two study team members created a codebook based on literature.^36,37^ After coding the first 10 interviews, the study team adjusted the codebook to reflect the current study’s specifications (such as adding codes to reflect interview questions).

Once coding was complete, final codes and themes were reviewed by the study’s principal investigator and co-investigator, and any disagreements were resolved through consensus with the coding team.

### Ethics

All study phases were approved by the local Institutional Review Board (approval number STUDY00003766). All study data was kept confidential. The results are presented in a manner that ensures no individual respondent can be identified. In Phase 1, CAB members were compensated with a $250 gift card for their time. In Phase 2, caregivers were compensated with a $25 gift card.

## Results

### Phase 1 Results

A total of eight individuals consented to be a part of the CAB in Phase 1. There were two social workers (25%), one nurse (12.5%), one pediatric psychologist, one physician, one clinical practice director, one psychosocial services director, and one community advocate from Momcology (a community-based patient advocacy and support organization). CAB members’ specific demographics are reported in Table 1.

**TABLE 1:**
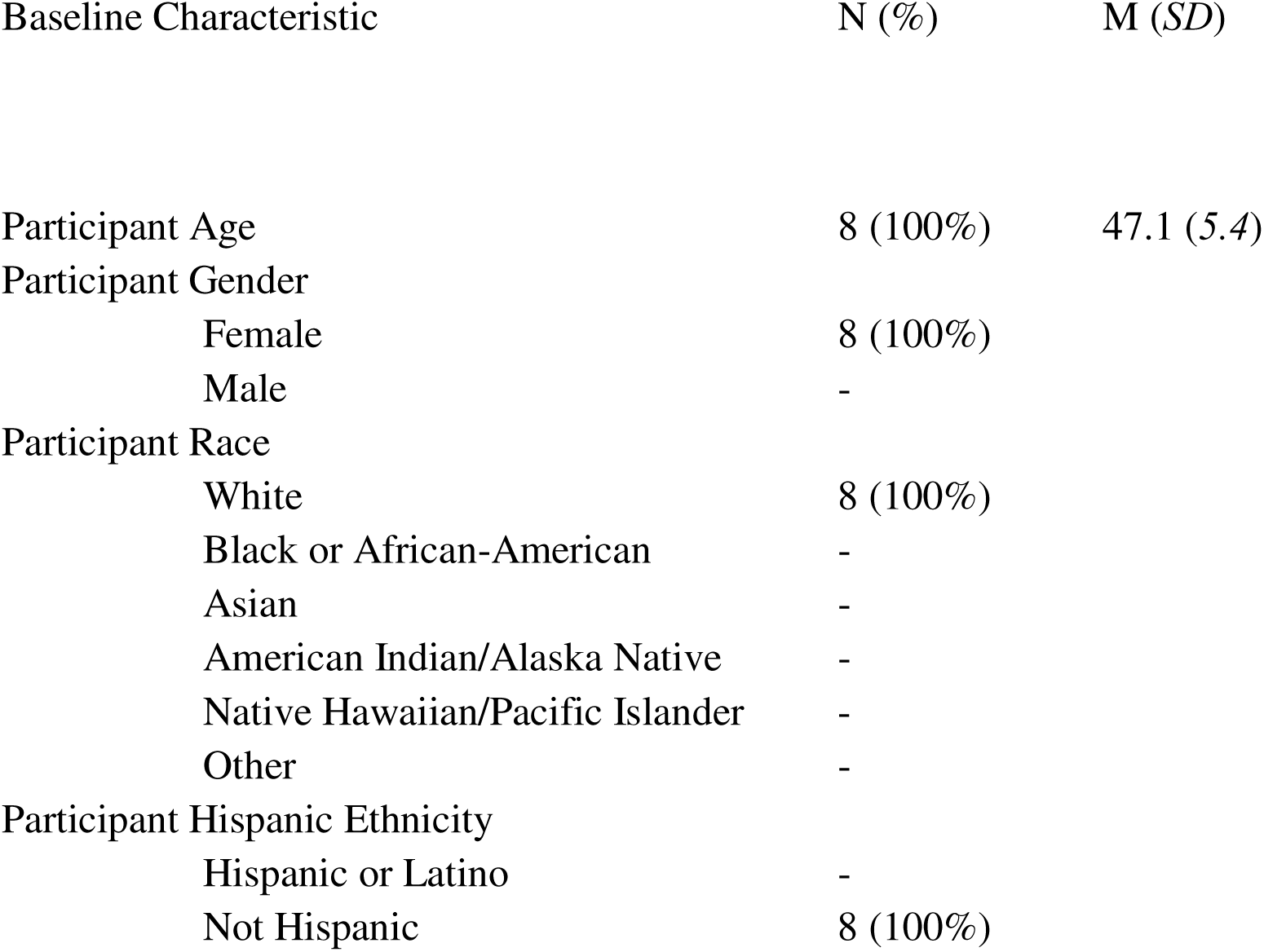
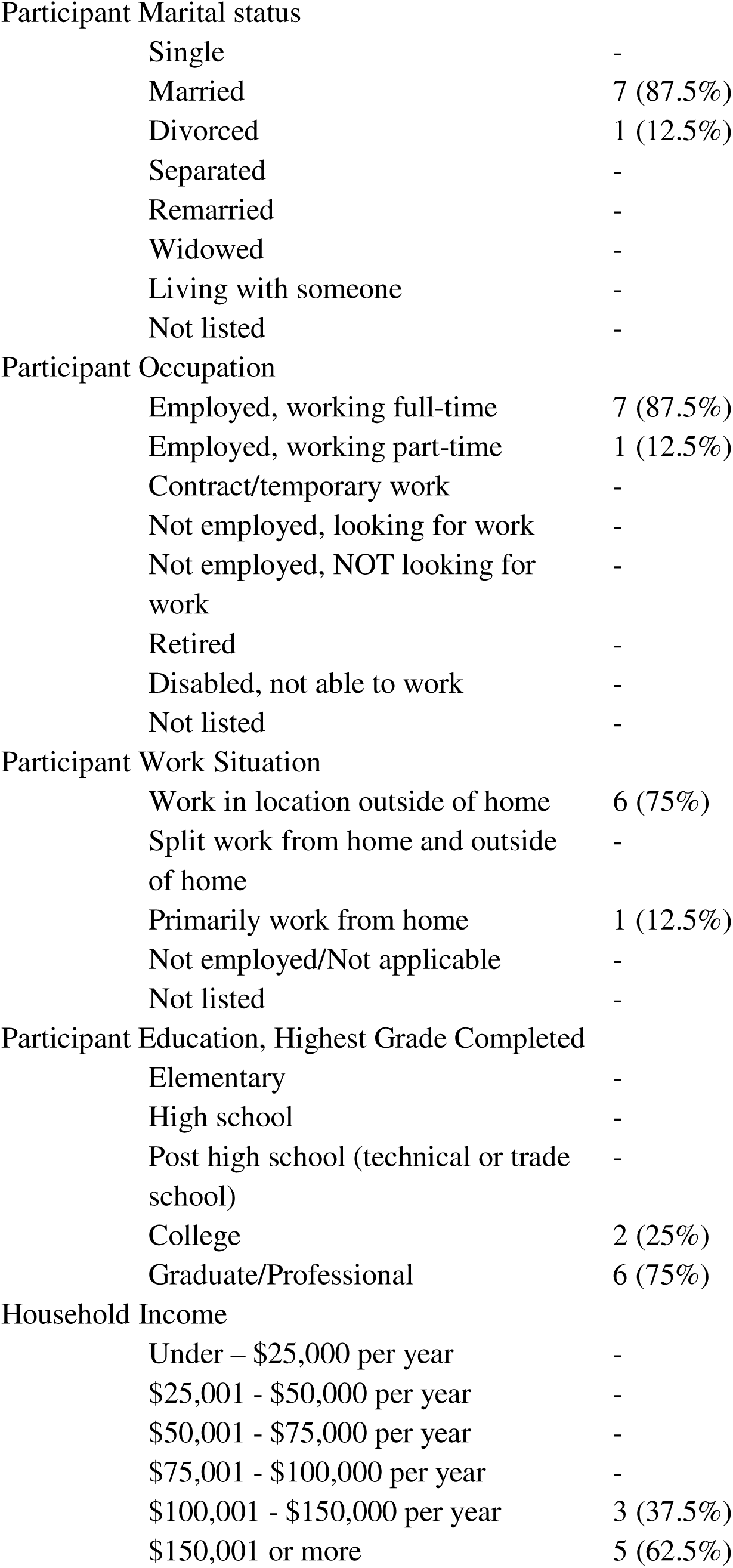
Phase 1: demographic characteristics of study population.

| Baseline Characteristic | N (%) | M (SD) |
| --- | --- | --- |
| Participant Age | 8 (100%) | 47.1 (5.4) |
| Participant Gender |  |  |
| Female | 8 (100%) |  |
| Male | - |  |
| Participant Race |  |  |
| White | 8 (100%) |  |
| Black or African-American | - |  |
| Asian | - |  |
| American Indian/Alaska Native | - |  |
| Native Hawaiian/Pacific Islander | - |  |
| Other | - |  |
| Participant Hispanic Ethnicity |  |  |
| Hispanic or Latino | - |  |
| Not Hispanic | 8 (100%) |  |

Participant Marital status
|  |  |
| --- | --- |
| Single | - |
| Married | 7 (87.5%) |
| Divorced | 1 (12.5%) |
| Separated | - |
| Remarried | - |
| Widowed | - |
| Living with someone | - |
| Not listed | - |

Participant Occupation
|  |  |
| --- | --- |
| Employed, working full-time | 7 (87.5%) |
| Employed, working part-time | 1 (12.5%) |
| Contract/temporary work | - |
| Not employed, looking for work | - |
| Not employed, NOT looking for work | - |
| Retired | - |
| Disabled, not able to work | - |
| Not listed | - |

Participant Work Situation
|  |  |
| --- | --- |
| Work in location outside of home | 6 (75%) |
| Split work from home and outside of home | - |
| Primarily work from home | 1 (12.5%) |
| Not employed/Not applicable | - |
| Not listed | - |

Participant Education, Highest Grade Completed
|  |  |
| --- | --- |
| Elementary | - |
| High school | - |
| Post high school (technical or trade school) | - |
| College | 2 (25%) |
| Graduate/Professional | 6 (75%) |

Household Income
|  |  |
| --- | --- |
| Under – \$25,000 per year | - |
| \$25,001 - \$50,000 per year | - |
| \$50,001 - \$75,000 per year | - |
| \$75,001 - \$100,000 per year | - |
| \$100,001 - \$150,000 per year | 3 (37.5%) |
| \$150,001 or more | 5 (62.5%) |

### Phase 1 Qualitative Data

Four major themes emerged from CAB sessions: 1) personalization and integration with the medical team to facilitate engagement, 2) language and technology accessibility as potential barriers to engagement, 3) simple and empathetic language to frame intervention, and 4) easy-to-navigate presentation. Using an iterative design approach, changes were made to the prototype based on CAB member suggestions. CAB exemplar quotes are shown in Table 2, and feedback implementation examples are shown in the **supplementary materials 1**. In the final CAB, members endorsed the VA, which indicated the prototype was ready for Phase 2.

**TABLE 2:** Exemplar quotes from CAB sessions.

| Suggestion | Exemplar Quotations |
| --- | --- |
| Framing content and language | CAB Member |
|  | “I think the prompting’s going to be key” |
|  | "You guys may want to consider phrasing it in a sort of a conversational directed at the person responding...trying to personalize it a little bit." |
|  | "We could say ‘it's good to hear that you're managing your stress well at this time.’ Something like that” |
| Perceived Facilitators to Engagement |  |
| Personalization | "Will it know your name?...It makes it feel personable as opposed to, you know, a bot." |
| Integration with medical team | “I think it being integrated is a great idea, because then it's like one less thing for the families to have to remember to tell their provider." |
| App analytics, reminders, and history | "The next time they go in, she could say “during our last chat, during our last conversation, I offered this resource. Did you have an opportunity to use it? Was it helpful to you?” |
| Potential Barriers to Engagement |  |
|  | "I'm also wondering availability in other languages and like what reading level it's being written at” |
|  | "One of the most mentioned things among our very stressed population that they cannot get to a mental health provider." |
|  | "And I think being very thoughtful about how often resources are updated, so I think we could potentially lose trust quickly if families are referred spaces that either aren't available or, um, you know." |
|  | "If families aren't using her because there is a report back or they're afraid that there that you know they want to use the resource...they're not asking their social worker intentionally, right? Umm, are we actually creating a barrier accidentally?" |
| **Suggestions on Presentation and Visuals** |  |
|  | "I think we should always be ending with 'do you need help with other resources?' And then it takes them back to those prompts. They might not understand to click that refresh button and then start the conversation over." |
|  | "looking at those three buttons, I think as a as a layman user, I would have a hard time identifying if I needed to, which one I needed to do." |

### Phase 2 Quantitative Results

In Phase 2, forty-one caregivers consented to participate; one withdrew following consent, resulting in a final sample of 40. Caregivers were primarily female (*n*=33, 82.6%), White (*n*=35, 87.5%), non-Hispanic, Latino, or Spanish (*n*=36, 90%), married (*n*=27, 67.5%), Christian (*n*=29, 72.5%), and had at least a high-school level education (*n*=13, 32.2%). Caregivers were split between employed working full time (*n*=13, 32.5%) and not employed and not looking for work (*n*=13, 32.5%), and most made between $25,001-50,000 a year. Children were mostly male (*n*=25, 62.5%), White (*n*=36, 90%), not Hispanic, Latino, or of Spanish origin (*n*=34, 85%), and were home-schooled (*n*=12, 30%). Demographics are reported in Table 3.

**TABLE 3.**
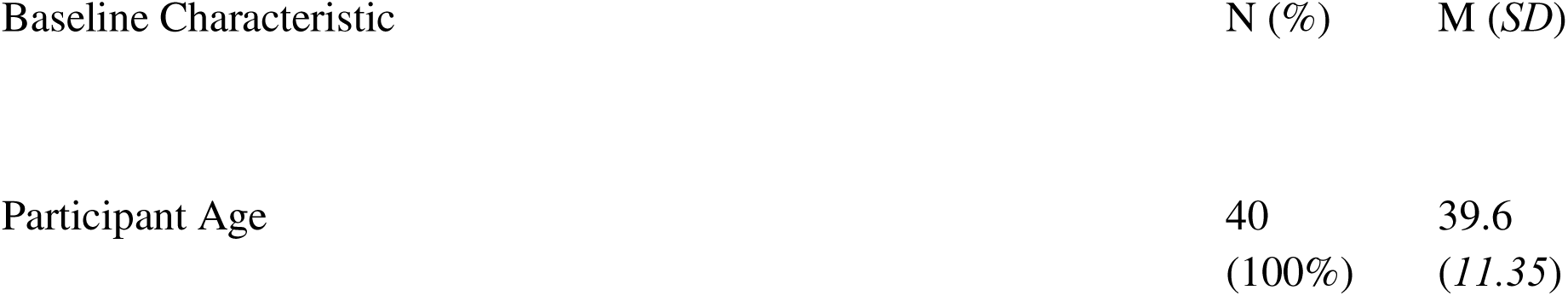
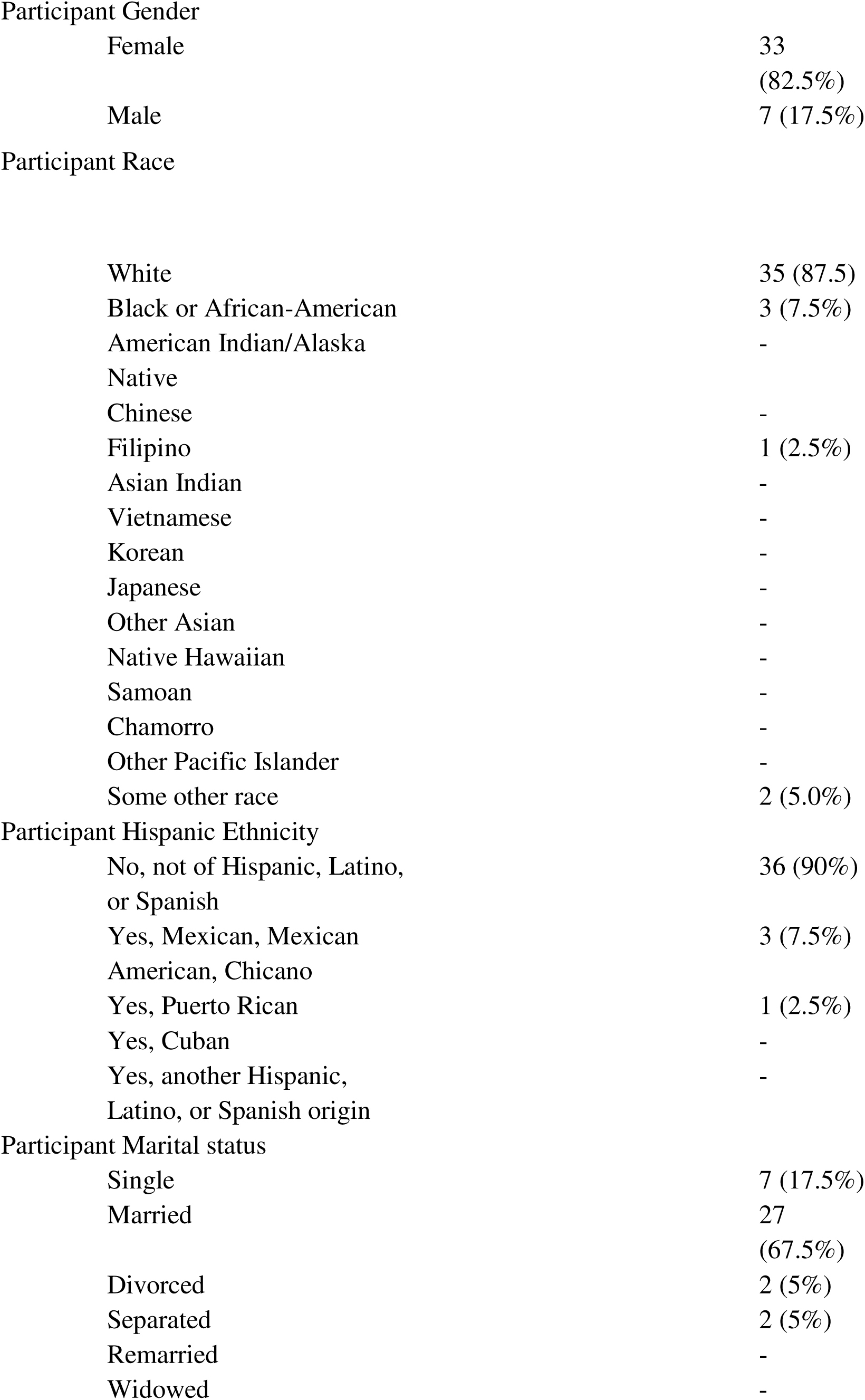
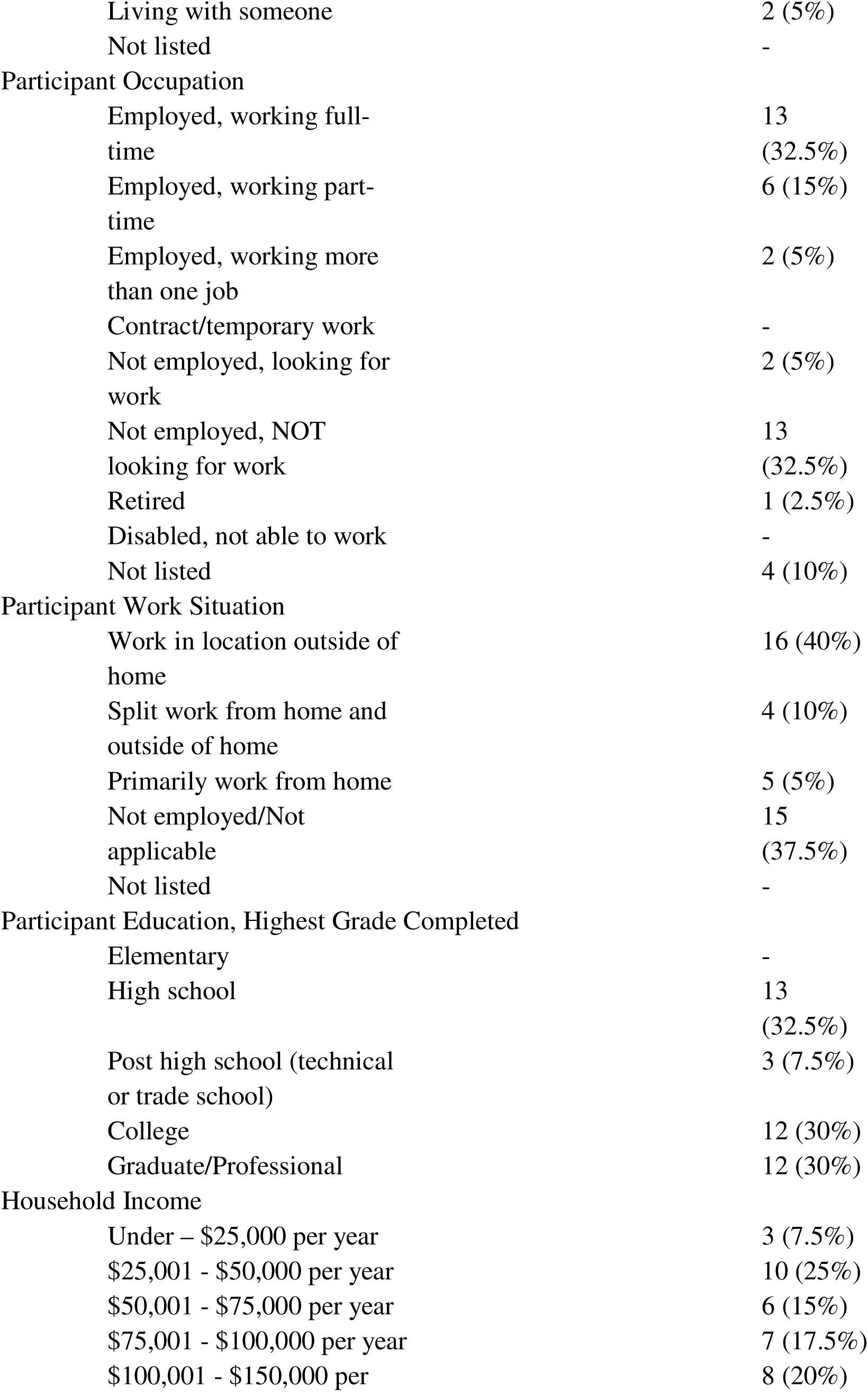
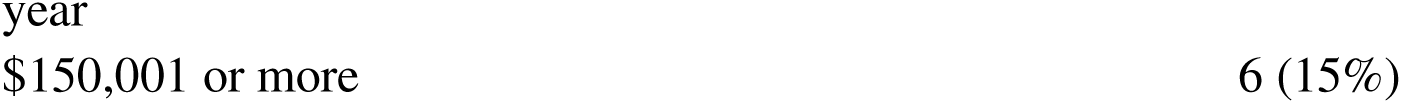
Phase 2: Demographic characteristics of study population.

### Usability

The mean SUS score in Phase 2 was 82.62 (*SD*=14.01), with a median of 82.50. Most caregivers reported the VA was easy to use (*n*=38, 95.0%), and others could learn it quickly (*n*=37, 94.9%; Figure 3). Bivariate correlations indicated lower socioeconomic status (SVI subcomponent) was associated with higher usability ratings, *r*(37) = 0.368, *p* = .025. Overall SVI showed a similar trend: greater vulnerability was associated with higher ratings, but this was only moderately significant *r*(37) = 0.317, *p* = .056. Correlations with individual items from the SUS revealed caregivers from a rural area were more likely to report they need the support of a technical person to use the VA *r*(40) = -0.382, *p* = .015 and felt less confident using the VA, *r*(39) = - 0.419, *p* = .008. Independent *t*-tests comparing binary SDoH variables and overall SUS scores showed Appalachian residents were somewhat more likely to rate the VA as more usable, compared to non-Appalachian residents, although this difference was only moderately significant *t*(38)= 1.64, *p*=.054.

**Figure 3.**
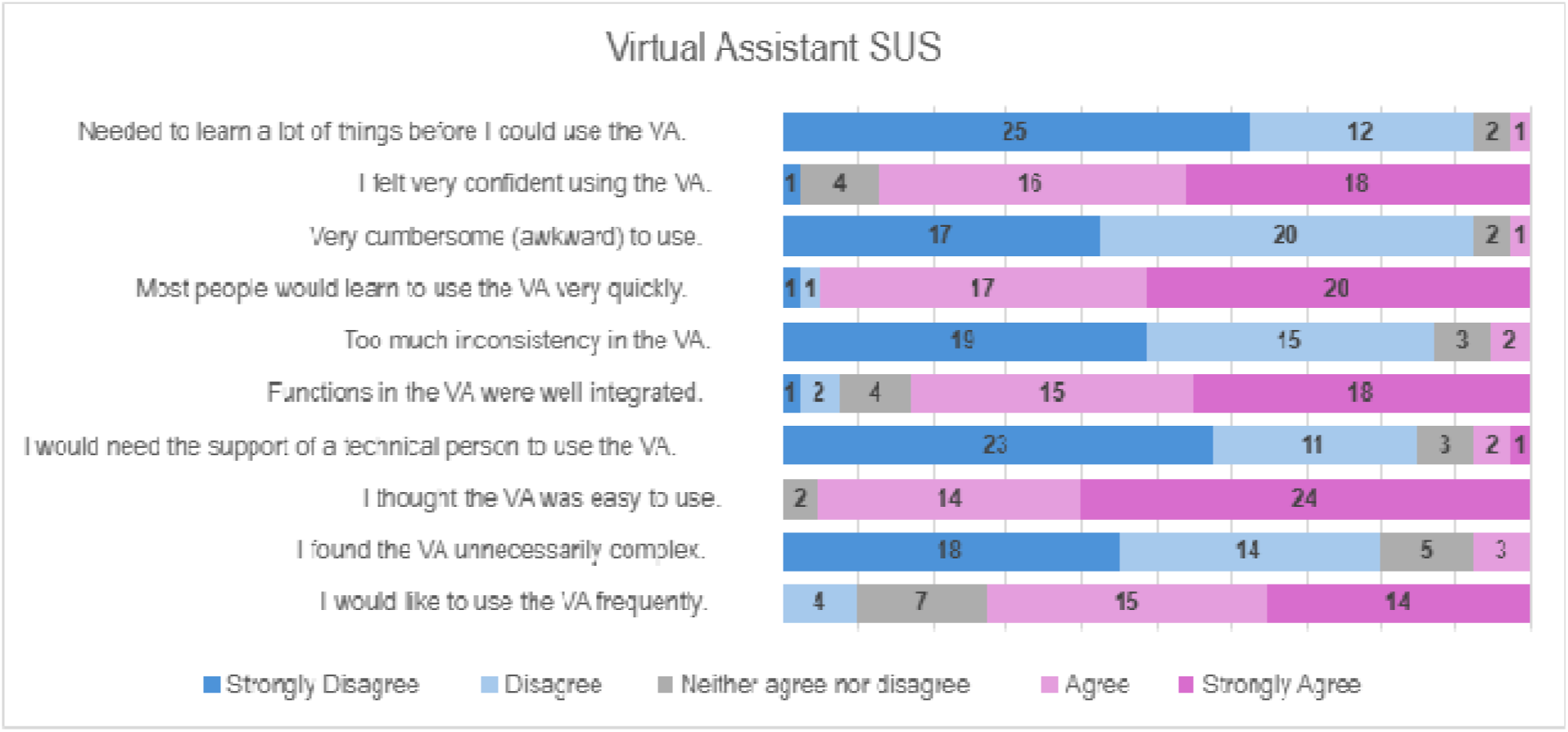
Virtual assistant SUS survey responses from caregivers. SUS: System Usability Scale.

### Caregiver satisfaction

Caregivers frequently rated the VA as easy to use (*n*=38, 95.0%) and good to use (*n*=38, 95.0%). Most also reported the VA motivated them to remain engaged during screening and in finding community resources (*n*=37, 92.5%; Figure 4). Bivariate correlations and independent *t*-tests between individual items on the Caregiver Satisfaction survey and SDoH variables were non-significant.

**Figure 4.**
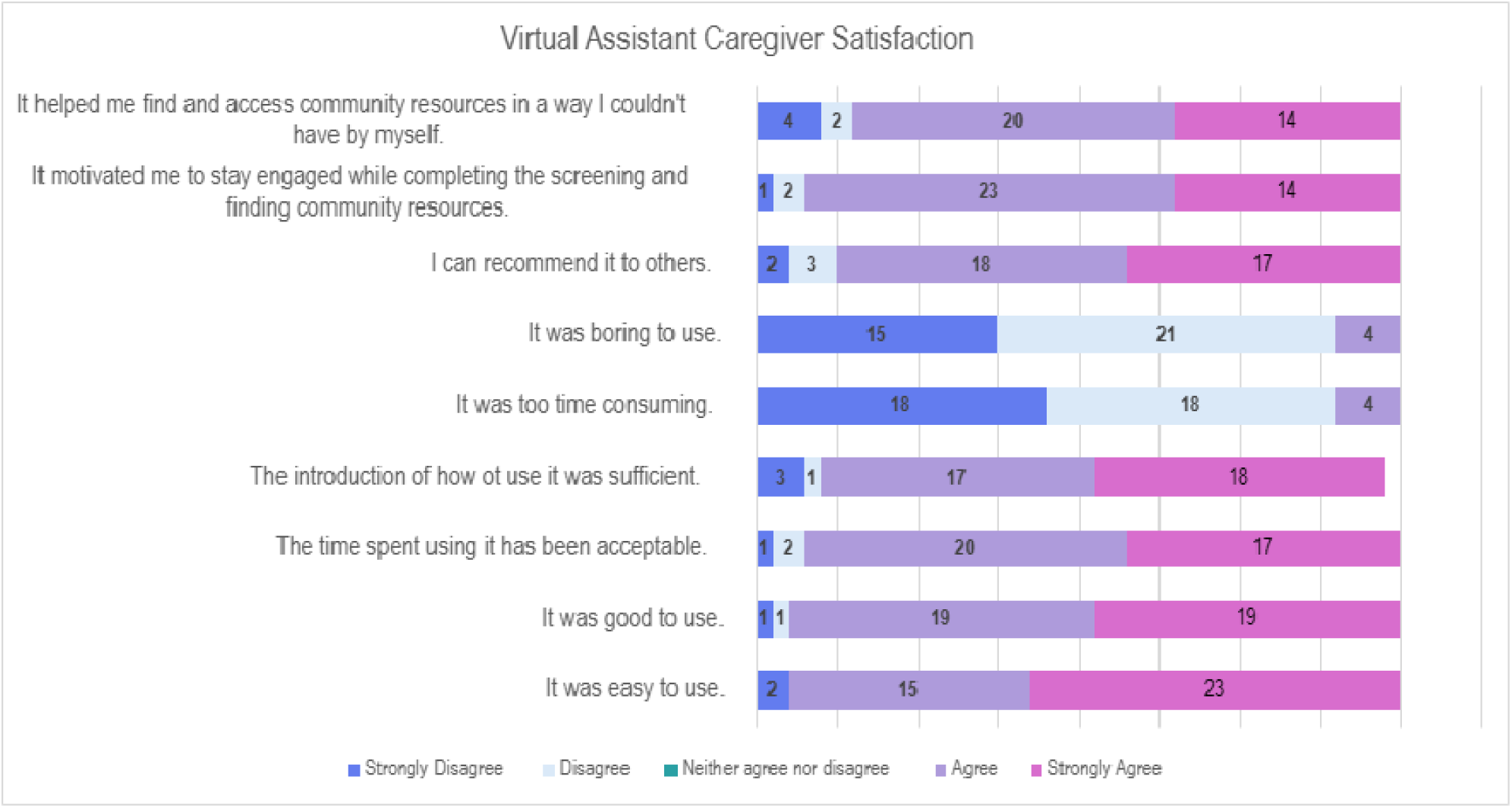
Caregiver Satisfaction survey responses.

### Reaction cards

Most caregivers reported positive experiences with the VA; accessibility (*n*=34, 83%), ease of use (*n*=34, 83%), and usefulness (*n*=32, 78%), represented the highest rated reactions (Figure 5). Bivariate correlations between Reaction Card items and SDoH were conducted. Caregivers from rural areas rated the VA as less collaborative *r*(40)=-403, *p*=.010, and less straightforward *r*(40) = -320, *p* = .044, compared to non-rural caregivers. Those residing in MUAs rated the VA as less attractive, *r*(40)= -489, *p*=.001, less empowering *r*(40) = -349, *p* = 027, more intimidating *r*(40)= .331, *p* = 037, and less useful *r*(40)= -.320, *p* = 044, than caregivers from medically served regions.

**Figure 5.**
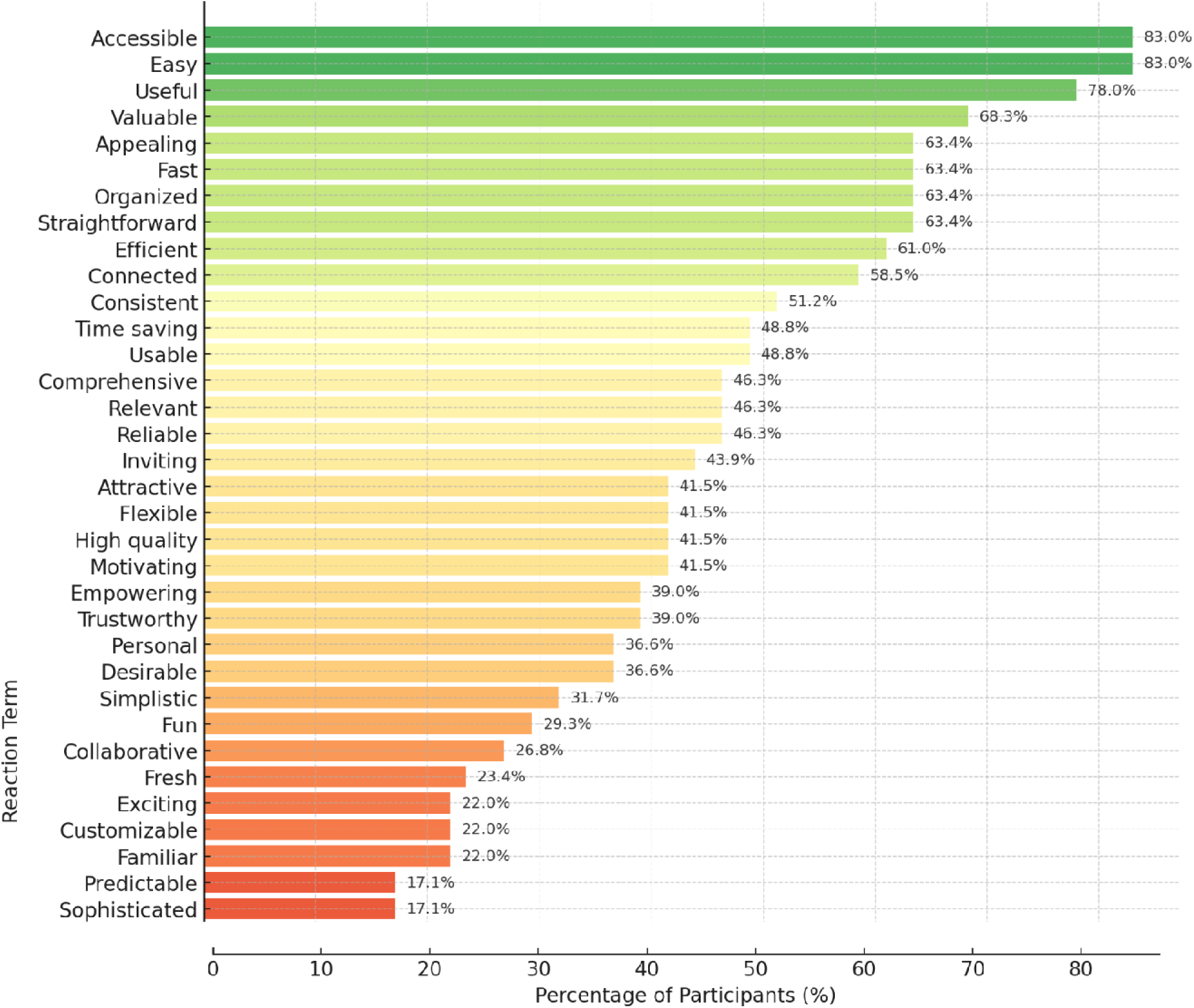
Reaction card results. <10% are excluded from the figure: Confusing (7.3%), Busy (4.9%), Complex(4.9%), Intimidating (4.9%), Not Valuable (4.9%), Gets in the way (2.4%), Inconsistent (2.4%), Overbearing (2.4%), Rigid (2.4%), Stimulating (2.4%), Time consuming (2.4%)

### Phase 2 Qualitative Results

Qualitative analyses of interviews uncovered three primary themes: 1) the acceptability of VA, attributed to its simple and effective design; 2) caregiver preferences regarding optimal VA usefulness; and 3) the applicability of VA resources to caregivers’ real-time needs.

#### VA acceptability through simple, effective design

Caregivers consistently reported that the VA prototype’s quality (i.e., personalization, clarity, presentation, attractiveness, functionality, and completeness) contributed to its ease of use and utility in addressing their current needs. The VA’s clear prompts, straightforward navigation, and appealing layout fostered trust and usability. For instance, one caregiver explained, “*the prompts were helpful. It makes it quick,*” while another remarked, “*the pictures are helpful by showing the movements*,” referring to the stress management module. Ease of navigation also enhanced engagement, as one caregiver noted, “*it was easy to navigate through. And um, pretty self-explanatory, you know what I mean? Which is good*,” while another commented, *”I think the straightforwardness of it is really nice and helpful.*” Caregivers further emphasized the value of personalization, with one individual suggesting, “*people have different aesthetics*,” which indicates potential for customizable features.

#### Caregiver preferences for optimal VA usefulness

Caregivers frequently offered suggestions regarding intervention timing, format, module preferences, non-technology preferences, and general recommendations for the VA. Many believed the VA would be most beneficial shortly after diagnosis. For instance, one caregiver remarked, “*it would have been very useful, especially at the very beginning of when he got-first got diagnosed*.” While the app format was generally favored by caregivers, with one caregiver stating, “*I think it’s just way easier to just have the app on your phone*,” some caregivers expressed disinterest in using an app, noting, “*I don’t want to download another app*.” Caregivers also advocated for integration with existing health systems, exemplified by comments such as, “*if it knew my schedule… if it’s connected to MyChart*,” and for broader content coverage, requesting, “*list some more resources or something. For the whole family, not just me.*”

#### Applicability of VA resources to caregivers’ real-time needs

Caregivers emphasized the accessibility, applicability, and relevancy of the VA, as well as whether they already possessed a system for addressing their social needs. For instance, caregivers largely perceived the VA as readily accessible. One caregiver stated, “*I don’t think that I would have any barriers. Um, all you really need is like a phone, internet connection um which we have, um a lot of places even if you don’t have Wi-Fi you can access it on.*” Caregivers also highlighted the VA’s potential to offer timely assistance during a cancer diagnosis, with one individual describing, “*it would be very helpful, for, um, patients and their families, especially their parents that are going through big transitions. Like, I went through*.” Furthermore, caregivers reflected on unmet needs and the VA’s capacity to fulfill them: “*we needed financial resources, I mean, so, had this been readily available to us, it would have been easier for us to access the resources that we needed*.” Moreover, caregivers underscored how the VA conserved time and effort compared to general searches. “*I liked it because I am a googler…but this is, a little more specific with the needs of someone in our situation, what we’re dealing with, so I can just easily go to the app instead of having the whole world of google to navigate through*.”

## Discussion

This study adapted and evaluated a VA aimed at improving social care within pediatric oncology, with specific focus on identifying unmet HRSN and mitigating toxic stress among caregivers. The key findings from our two-phased, mixed-methods approach demonstrate the promise of digital health technologies in providing accessible social care support to this vulnerable population. The co-design process with a CAB proved invaluable in refining the VA, and subsequent testing with caregivers revealed high usability and acceptability. The iterative feedback from the CAB, composed of multidisciplinary experts and a community advocate, was instrumental in shaping a VA that was perceived as personalized, accessible, and empathetic. This aligns with previous research emphasizing the importance of co-design in creating digital health tools that are engaging and meet the specific needs of end-users.^38–43^

Co-design in phase 1 with the CAB directly shaped the prototype tested in Phase 2. CAB priorities, (empathetic, plain-language prompts; simple, guided navigation with explicit “next-step” options; visible pathways to the care team; and clear resource provenance) were implemented before user testing. In Phase 2, these design choices corresponded with high SUS scores and desirability ratings (“easy,” “accessible,” “useful”), and caregivers repeatedly cited straightforward navigation and concise prompts as reasons for trust and engagement. Risks anticipated by the CAB (language/technology access and resource freshness) aligned with subgroup patterns: rural caregivers more often reported needing technical support and lower confidence, and those in MUAs perceived the VA as less attractive, less empowering, and more intimidating.

The quantitative results from Phase 2 supports the co-design process. The mean SUS score of 82.62 is well above the industry average of 68, indicating a highly usable system. Caregivers overwhelmingly found the VA easy to use and were satisfied with its functionality. These findings are consistent with other studies that have shown high usability and acceptability of digital health tools among caregivers of children with cancer.^43–45^ An interesting finding was the association between higher social vulnerability and higher usability ratings. Caregivers with a lower socioeconomic status and those residing in Appalachian regions rated the VA as more usable. This suggests that the VA may be particularly beneficial for underserved populations who often face greater barriers to accessing traditional support services. Digital health solutions like our VA have the potential to bridge these gaps by providing readily accessible resources.^15,44,46^ Furthermore, the themes that emerged from qualitative results (acceptability through simple and effective design, preferences for optimal usefulness, and the real-time applicability of resources) further validate the quantitative data. Caregivers appreciated the VA’s straightforward navigation and personalized content, emphasizing its potential to be a valuable resource, particularly at the time of diagnosis when families are often overwhelmed. The desire for integration with existing electronic health records, such as patient portals, reflects a broader trend in digital health towards seamless and integrated patient care.^16,47,48^ However, our findings also point to a potential digital divide. Caregivers from rural areas and MUAs reported needing more technical support and found the VA less collaborative and straightforward. This is a critical consideration for the implementation and dissemination of digital health interventions, as it highlights the need for additional support and training for certain user groups to ensure equitable access and usability.^46,49^

### Interpretation and Implications for Social Care

Our findings add to a growing literature showing that digital health tools can support the social and mental health needs of children and families, and that VAs, in particular, are a scalable, user-friendly modality for ongoing communication and guidance.^13^ In pediatric oncology, where families navigate intense psychosocial and financial pressures, our results suggest that a co-designed VA can do more than screen: it can organize information, reduce cognitive load at critical moments, and provide a consistent point of entry into social care. By centering HRSN alongside stress-coping content, the VA operationalizes a holistic model in which identification, brief support, and connection to services occur within the same interaction. Given the well-established links between caregiver stress and child outcomes, this coupling of HRSN screening with toxic-stress mitigation is a meaningful advance in a context where targeted interventions remain limited.^50^

The pattern that families with higher social vulnerability rated the VA as highly usable implies that well-designed conversational pathways may lower common barriers to help-seeking (time, navigation burden, and uncertainty about trustworthy resources) thereby shortening the path from identified need to referral and uptake. At the same time, the experience of rural and MUA caregivers in our study mirrors the broader digital divide: variable connectivity, device access, and digital confidence can dampen perceived collaboration and empowerment. The implication is not that VAs are unsuitable for these settings, but that their benefit depends on equity-oriented implementation, e.g., low-literacy and multilingual content, audio/visual options, low-bandwidth performance, clear routes to human support, and reliable, up-to-date community resource curation. Taken together, the literature and our data point to a practical role for VAs as front doors to social care in pediatric oncology (tools that can surface unmet HRSN, deliver just-in-time stress support, and close loops with the care team) provided deployment strategies directly confront access and trust barriers in underserved contexts.^46^

### Limitations and Future Directions

This study has several limitations that should be considered. The Phase 1 CAB was a small and homogenous group of predominantly White, female, and individuals with high education. This lack of diversity may limit the generalizability of the initial design feedback. The Phase 2 sample of caregivers, while larger, was also primarily White and non-Hispanic. Future research should aim to recruit more diverse participants to ensure the VA is acceptable and usable across different racial, ethnic, and socioeconomic groups. The study was conducted at a single, large Midwestern children’s hospital, which may limit the external validity of our findings. The cross-sectional design of Phase 2 provides a snapshot of usability and acceptability but does not allow for an assessment of the VA’s long-term impact on caregiver stress or health outcomes. Finally, while the VA was highly rated, the study did not include a control group to compare its effectiveness against traditional social care support.

A longitudinal study with a more diverse sample of caregivers is needed to evaluate the long-term effectiveness of the VA in reducing toxic stress, improving coping, and addressing unmet social needs. A randomized controlled trial would be beneficial to compare the VA with standard care and determine its clinical and cost-effectiveness. Further refinement of the VA should incorporate the feedback from caregivers, including the desire for greater personalization and integration with electronic health records. Addressing the digital divide identified in our study is also a critical next step. This could involve providing devices and internet access to families in need, as well as offering tailored training and technical support. Exploring the use of the VA in other pediatric chronic illness populations facing similar social and emotional challenges is another promising avenue for future research.

## Conclusions

This study provides strong evidence for the feasibility, acceptability, and usability of a virtual assistant designed to provide social care support to caregivers of children with cancer. The stakeholder-informed co-design process was critical to its success, resulting in a tool that was highly rated by end-users. While the findings are promising, particularly for reaching underserved populations, they also highlight the importance of addressing the digital divide to ensure equitable access to digital health interventions. The VA developed in this study has the potential to be a valuable resource for families navigating the challenges of pediatric cancer, and with further research and development, it could be scaled to support a broader population of children with chronic illnesses and their families.

## Data Availability

All data produced in the present study are available upon reasonable request to the authors

## Acknowledgements

We are thankful to FindHelp.org for the research fellowship (Sezgin), and to MEMOTEXT for their technical support on chatbot UI elements.

