## Supplemental materials for "Supporting Families of Children with Cancer: Development and Evaluation of a Virtual Assistant for Social Care"

### **Supplementary Material 1**

| **Feedback from CAB** | **Implementation** |  |
| --- | --- | --- |
| Including a crisis line in managing stress module (CAB 3) | A link to a crisis line was added when users indicated high stress | 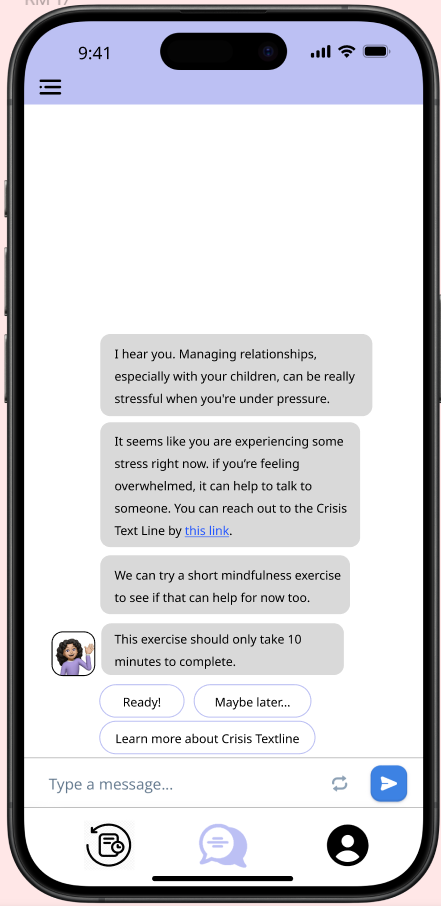 |
| Opportunity for users to provide feedback on helpfulness of resources | Study team incorporated a prompt for feedback after every conversation | 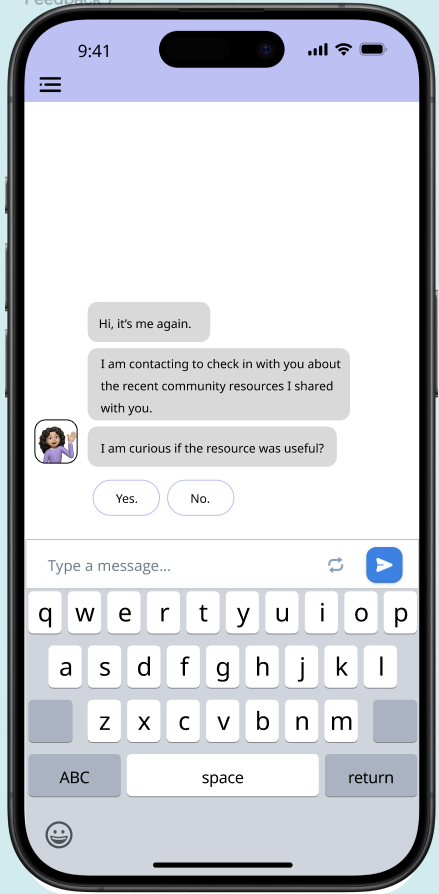 |
| VA should offer stress management tool even when screening results in “low stress” | Mindfulness and breathing exercises were added that appeared regardless of level of stress | 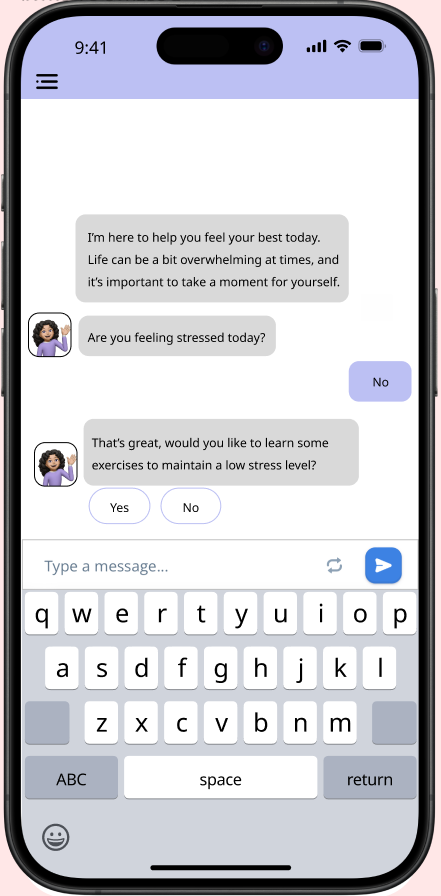 |
| Creating an intuitive way to decide what to do next when finding resources | Study team added prompts when opening the finding resources module to suggest possible resources for users | 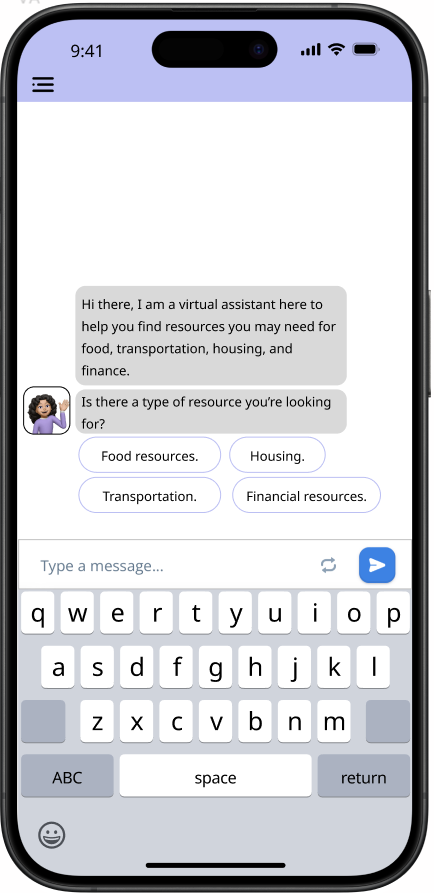 |
| Keeping a report of what the user has searched before so they can reference back | A history tab was added to the prototype home page, as well a favorite-ing feature | 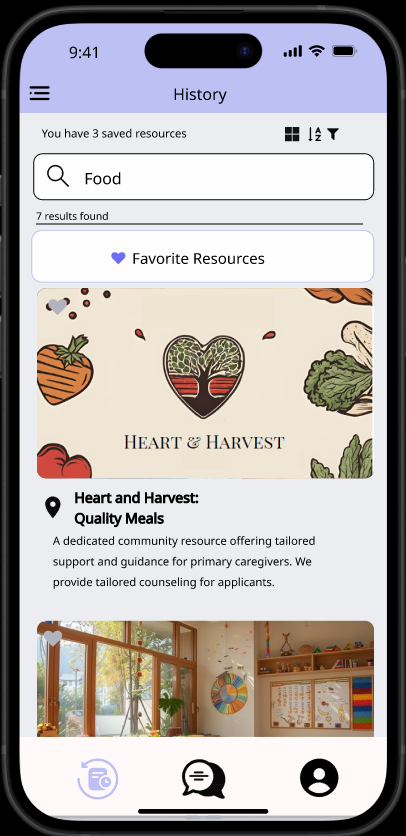 |
